# Coronavirus Disease 2019 (COVID-19) Vaccine Boosting in Persons Already Protected by Natural or Vaccine-Induced Immunity

**DOI:** 10.1101/2022.02.10.22270744

**Authors:** Nabin K. Shrestha, Priyanka Shrestha, Patrick C. Burke, Amy S. Nowacki, Paul Terpeluk, Steven M. Gordon

## Abstract

**Background:** The purpose of this study was to evaluate whether boosting healthcare personnel, already reasonably protected by prior infection or vaccination, with a vaccine developed for an earlier variant of COVID-19 protects against the Omicron variant.

**Methods:** Employees of Cleveland Clinic who were previously infected with or vaccinated against COVID-19, and were working in Ohio the day the Omicron variant was declared a variant of concern, were included. The cumulative incidence of COVID-19 was examined over two months during an Omicron variant surge. Protection provided by boosting (analyzed as a time-dependent covariate) was evaluated using Cox proportional hazards regression. Analyses were adjusted for time since proximate overt immunologic challenge (POIC) as a time-dependent covariate.

**Results:** Among 39 766 employees, 8037 (20%) previously infected and the remaining previously vaccinated, COVID-19 occurred in 6230 (16%) during the study. Risk of COVID-19 increased with time since POIC. In multivariable analysis, boosting was independently associated with lower risk of COVID-19 among those with vaccine-induced immunity (HR, .43; 95% CI, .41-.46) as well as those with natural immunity (HR, .66; 95% CI, .58-.76). Among those with natural immunity, receiving 2 compared to 1 dose of vaccine was associated with higher risk of COVID-19 (HR, 1.54; 95% CI, 1.21-1.97).

**Conclusions:** Administering a COVID-19 vaccine not designed for the Omicron variant, 6 months or more after prior infection or vaccination, protects against Omicron variant infection in both previously infected and previously vaccinated individuals. There is no evidence of an advantage to administering more than 1 dose of vaccine to previously infected persons.

**Summary:** Among 39 766 Cleveland Clinic employees already protected by prior infection or vaccination, vaccine boosting after 6 months was associated with significantly lower risk of COVID-19. After COVID-19 infection, there was no advantage to more than one dose of vaccine.

## INTRODUCTION

By the time the Delta variant of severe acute respiratory syndrome-associated coronavirus 2 (SARS-CoV-2) became the predominant strain in the United States, it was already several months after the majority of early vaccine recipients had received their vaccines. A small proportion of vaccinated individuals experienced breakthrough infections. Studies suggested that waning immunity may at least partially explain the breakthrough infections, and vaccine boosters began to be administered in some resource-rich countries, with an expectation that waning vaccine-induced immunity might be boosted by an additional dose of vaccine. Nationwide studies from Israel showed that a booster dose did indeed provide significant protection against coronavirus disease 2019 (COVID-19) [1–3].

The Omicron variant was first identified in South Africa in mid-November 2021, and was declared a variant of concern (VOC) by the World Health Organization on 26 November 2021. This was more contagious than the Delta variant [4], and became the predominant strain in the United States within 3 weeks of the first positive test within the USA on 29 November 2021. Public health officials strongly recommended that all get a vaccine booster to control its spread. However, by this time it was known that this variant had a large number of mutations, including several on the spike protein itself [5,6], the target of COVID-19 vaccines, raising the possibility that vaccine effectiveness against the new variant might be seriously compromised. Additionally, although very rare previously, with the emergence of the Omicron variant, a surprisingly large proportion of previously infected individuals experienced reinfections [7,8], and breakthrough infections in vaccinated individuals were also very common [9,10], including among those in our own practice who had received a vaccine booster. These observations raised questions about the utility of boosting those with pre-existing natural or vaccine-induced immunity, with a vaccine not specifically designed for the new variant.

The purpose of this study was to evaluate whether boosting individuals already reasonably protected by prior infection or vaccination, with a vaccine developed for an earlier variant of COVID-19, protects against infection with the Omicron variant.

## METHODS

### Study design

This was a retrospective cohort study conducted at the Cleveland Clinic Health System in Ohio, United States. The study was approved by the Cleveland Clinic Institutional Review Board as exempt research (IRB no. 21-1163). A waiver of informed consent and waiver of HIPAA authorization were approved to allow access to de-identified health information by the research team.

### Setting

Beginning in March 2020, all employees at Cleveland Clinic with a positive SARS-CoV-2 test were interviewed and symptoms monitored remotely by Occupational Health while the employees were isolated at home. Voluntary vaccination for COVID-19 began on 16 December 2020. Most employees were vaccinated with two doses of an mRNA vaccine, either the Pfizer-BioNTech vaccine or the Moderna vaccine. Individuals began receiving booster vaccine of their own accord in August 2021, and the healthcare system officially began offering vaccine boosters on 5 October 2021.

### Participants

All employees of the Cleveland Clinic Health System working in Ohio as of December 16, 2020, the day employee COVID-19 vaccination was started, were screened for inclusion in the study. Those who were protected by natural or vaccine-induced immunity and remained in employment as of 26 November 2021, the day the Omicron variant was declared a VOC, were included. An individual was considered protected by natural immunity 14 days after testing positive for COVID-19 by a nucleic acid amplification test (NAAT). If not previously infected, a person was considered protected by vaccine-induced immunity 14 days after receipt of the second vaccine dose of an mRNA vaccine. By only screening individuals who had been in employment since vaccination started almost a year prior to the study start date, we could ensure accurate prior vaccination data and be reasonably assured of not having missed a prior COVID-19 diagnosis, at least up to a year.

### Variables

A vaccine booster was defined as at least 1 dose of any COVID-19 vaccine at least 90 days following COVID-19 infection for those with natural immunity (i.e. those previously infected), or a third dose of a COVID-19 vaccine at least 90 days following the second dose of an mRNA COVID-19 vaccine for those with vaccine-induced immunity (i.e. those not previously infected). Individuals were considered boosted 7 days after receipt of a qualifying vaccine booster. Covariates collected were age, aggregated job title (to maintain anonymity for rare job titles), job location, and job type categorization into patient-facing or non-patient facing, as described in an earlier study [11]. Protected health information identifiers were not included in the extracted data, and institutional data governance rules related to employee data limited our ability to supplement our dataset with additional clinical variables.

### Outcome

The primary study outcome was time to COVID-19, the latter defined as a positive NAAT for SARS-CoV-2 any time after 26 November 2021, the study start date. The date of infection for any episode of COVID-19 was the date of the first positive test for that episode of illness. Subsequent positive tests within 90 days were considered part of the same episode of illness. The health system never had a requirement for systematic asymptomatic employee test screening. Most of the positive tests would have been tests done to evaluate suspicious symptoms or as part of quarantine and return-to-work testing of employees exposed to patients with COVID-19. A small proportion would have been tests done as part of pre-operative or pre-procedural screening.

Time to symptomatic COVID-19 and time to hospitalization for COVID-19 were planned as secondary outcomes. Unfortunately, employee health monitoring processes had to be stopped about 21 days after the study start date due to inability to keep up with a very large number of cases.

### Statistical analysis

Boosting status of a study subject could change on any day of the study. To account for this, “boosting” was treated as a time-dependent covariate whose value changed from “non-boosted” to “boosted” 7 days after receipt of a vaccine booster. Additionally, risk of COVID-19 would be influenced by prior exposure to the causative pathogen or its antigens, and on the time elapsed since the last such encounter. To adjust for this, we defined the proximate overt immunologic challenge (POIC) as the most recent exposure to SARS-CoV-2 by infection or vaccination. Since this could also change on any day for any study subject, time (in days) since POIC, was a time-dependent covariate.

A Simon-Makuch hazard plot [12] was created to compare the cumulative incidence of COVID-19 among subjects classified by type of immunity (natural or vaccine-induced) and boosting status (boosted or non-boosted, as a time-dependent covariate). Employees who had not developed COVID-19 were censored at the end of the study follow-up period (28 January 2022). Those whose employment was terminated during the study period before they had COVID-19 (216 subjects) were censored on the date of termination of employment. Curves for the non-boosted were based on data for as long as the booster status remained “non-boosted”. Curves for the boosted were based on data from the date the booster status changed to “boosted”, until the study end date.

To evaluate the effect of time since POIC on risk of COVID-19, Simon-Makuch hazard plots comparing the cumulative incidence of COVID-19 for groups stratified by time since POIC, were plotted separately for those with natural immunity and those with vaccine-induced immunity. Subjects were censored on the date they were terminated as in the primary analysis. Time since POIC could change for any subject any day over the course of the study, and subjects moved from one subgroup to another as they crossed the limits of the time groups. Receipt of a COVID-19 vaccine was considered an immunologic challenge and time since POIC changed accordingly on the date of receipt of vaccine.

Among those with natural immunity, the effect of timing of a booster, and the effect of number of doses of vaccine, on risk of COVID-19, were examined in separate Simon-Makuch hazard plots. For the former, groupings were based on time since prior infection and boosting as separate time-dependent covariates. For the latter, the number of vaccine doses was calculated as a time-dependent covariate (as it could change for any subject on any day of the study).

Multivariable Cox proportional hazards regression models were fitted to examine associations of various variables with time to COVID-19, separately for those with natural immunity and those with vaccine-induced immunity. Where included, boosting, time since POIC, time since prior infection, and number of vaccine doses were included as time-dependent covariates [13]. These models were also explored in subsets divided by time since prior infection (for those with natural immunity) and time since second vaccine dose (for those with vaccine-induced immunity).

The analysis was performed by N. K. S. and A. S. N. using the *survival* package and R version 4.1.2 (R Foundation for Statistical Computing) [13–15].

## RESULTS

Of 39 766 employees included in the study, 8037 (20%) were previously protected by natural immunity and 31 729 (80%) by vaccine-induced immunity. By the end of the study, 26 176 (66%) were boosted. Altogether, 6230 employees (16%) acquired COVID-19 during the 9 weeks of the study.

### Baseline characteristics

Table 1 shows the characteristics of subjects grouped by type of immunity protection at the start of the study. At the start of the study, the median duration since achieving protected status was 331 days (IQR 228-363 days) for those protected by natural immunity, and 275 days (IQR 228-283 days) for those protected by vaccine-induced immunity.

**Table 1.**
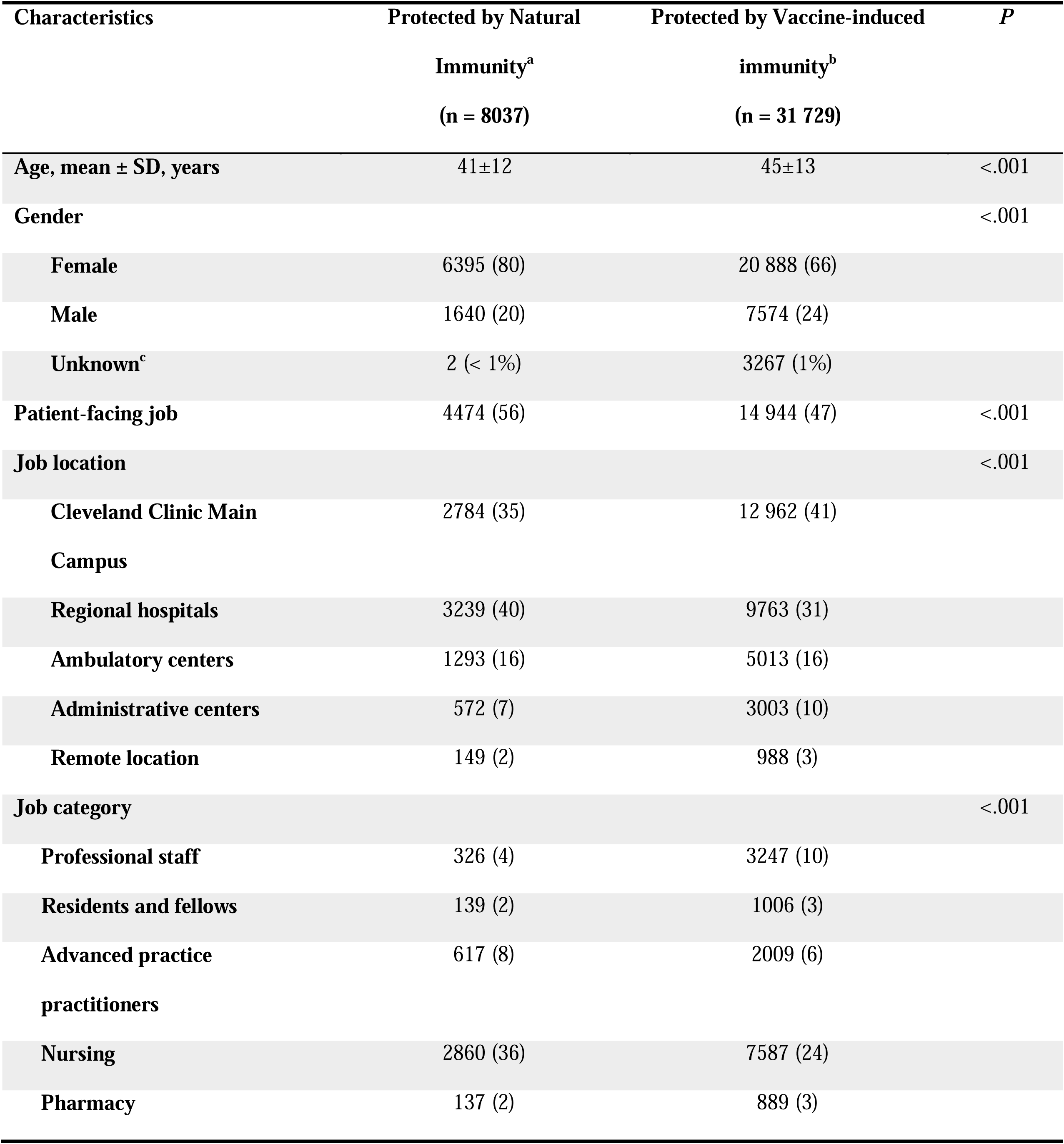
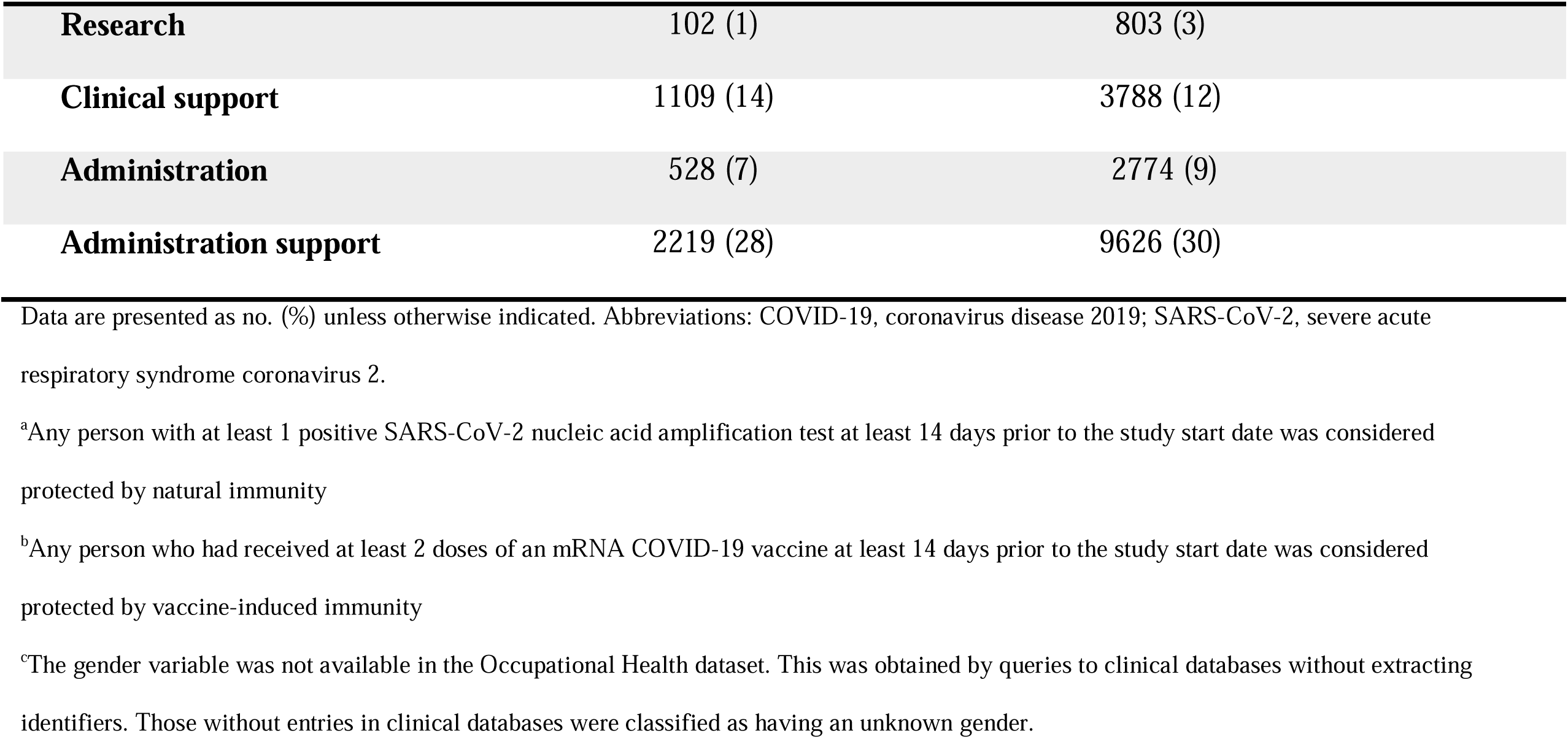
Study Subject Characteristics Compared by Type of Prior Protection.

| Characteristics | Protected by Natural<br>Immunity<sup>a</sup><br>(n = 8037) | Protected by Vaccine-induced<br>immunity<sup>b</sup><br>(n = 31 729) | *P* |
| --- | --- | --- | --- |
| Age, mean ± SD, years | 41±12 | 45±13 | <.001 |
| Gender |  |  | <.001 |
| Female | 6395 (80) | 20 888 (66) |  |
| Male | 1640 (20) | 7574 (24) |  |
| Unknown<sup>c</sup> | 2 (< 1%) | 3267 (1%) |  |
| Patient-facing job | 4474 (56) | 14 944 (47) | <.001 |
| Job location |  |  | <.001 |
| Cleveland Clinic Main<br>Campus | 2784 (35) | 12 962 (41) |  |
| Regional hospitals | 3239 (40) | 9763 (31) |  |
| Ambulatory centers | 1293 (16) | 5013 (16) |  |
| Administrative centers | 572 (7) | 3003 (10) |  |
| Remote location | 149 (2) | 988 (3) |  |
| Job category |  |  | <.001 |
| Professional staff | 326 (4) | 3247 (10) |  |
| Residents and fellows | 139 (2) | 1006 (3) |  |
| Advanced practice<br>practitioners | 617 (8) | 2009 (6) |  |
| Nursing | 2860 (36) | 7587 (24) |  |
| Pharmacy | 137 (2) | 889 (3) |  |

|  |  |  |
| --- | --- | --- |
| **Research** | 102 (1) | 803 (3) |
| **Clinical support** | 1109 (14) | 3788 (12) |
| **Administration** | 528 (7) | 2774 (9) |
| **Administration support** | 2219 (28) | 9626 (30) |

Data are presented as no. (%) unless otherwise indicated. Abbreviations: COVID-19, coronavirus disease 2019; SARS-CoV-2, severe acute respiratory syndrome coronavirus 2.
<sup>a</sup>Any person with at least 1 positive SARS-CoV-2 nucleic acid amplification test at least 14 days prior to the study start date was considered protected by natural immunity
<sup>b</sup>Any person who had received at least 2 doses of an mRNA COVID-19 vaccine at least 14 days prior to the study start date was considered protected by vaccine-induced immunity
<sup>c</sup>The gender variable was not available in the Occupational Health dataset. This was obtained by queries to clinical databases without extracting identifiers. Those without entries in clinical databases were classified as having an unknown gender.

Table 2 shows the characteristics of subjects grouped by their boosting status by the end of the study. For those boosted, the median time to being boosted was 16 days prior to the study start date (IQR-38 to 6 days).

**Table 2.**
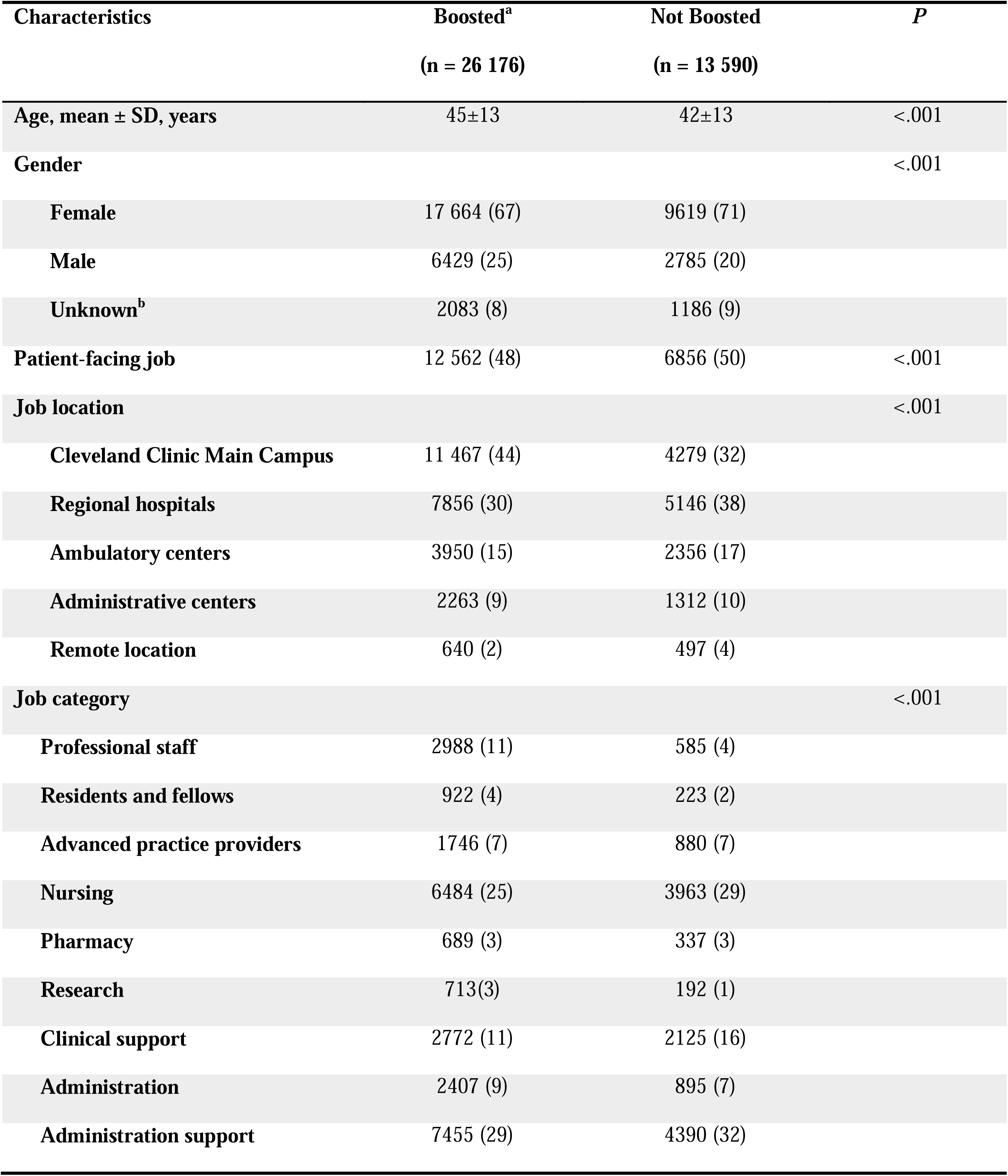
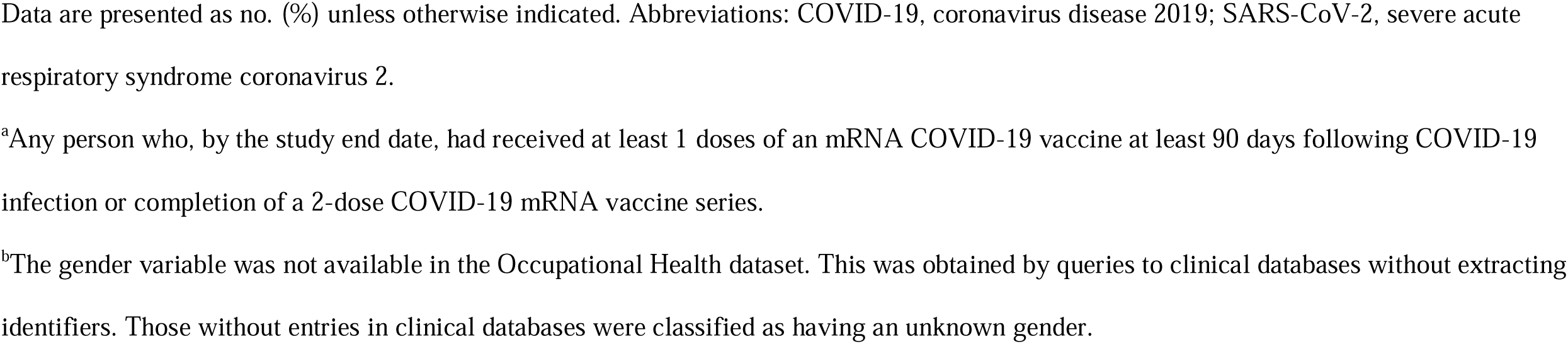
Study Subject Characteristics Compared by Booster Receipt Status by the End of the Study.

| **Characteristics** | **Boosted<sup>a</sup>** | **Not Boosted** | ***P*** |
| --- | --- | --- | --- |
|  | **(n = 26 176)** | **(n = 13 590)** |  |
| **Age, mean ± SD, years** | 45±13 | 42±13 | <.001 |
| **Gender** |  |  | <.001 |
| **Female** | 17 664 (67) | 9619 (71) |  |
| **Male** | 6429 (25) | 2785 (20) |  |
| **Unknown<sup>b</sup>** | 2083 (8) | 1186 (9) |  |
| **Patient-facing job** | 12 562 (48) | 6856 (50) | <.001 |
| **Job location** |  |  | <.001 |
| **Cleveland Clinic Main Campus** | 11 467 (44) | 4279 (32) |  |
| **Regional hospitals** | 7856 (30) | 5146 (38) |  |
| **Ambulatory centers** | 3950 (15) | 2356 (17) |  |
| **Administrative centers** | 2263 (9) | 1312 (10) |  |
| **Remote location** | 640 (2) | 497 (4) |  |
| **Job category** |  |  | <.001 |
| **Professional staff** | 2988 (11) | 585 (4) |  |
| **Residents and fellows** | 922 (4) | 223 (2) |  |
| **Advanced practice providers** | 1746 (7) | 880 (7) |  |
| **Nursing** | 6484 (25) | 3963 (29) |  |
| **Pharmacy** | 689 (3) | 337 (3) |  |
| **Research** | 713(3) | 192 (1) |  |
| **Clinical support** | 2772 (11) | 2125 (16) |  |
| **Administration** | 2407 (9) | 895 (7) |  |
| **Administration support** | 7455 (29) | 4390 (32) |  |

Data are presented as no. (%) unless otherwise indicated. Abbreviations: COVID-19, coronavirus disease 2019; SARS-CoV-2, severe acute respiratory syndrome coronavirus 2.
<sup>a</sup>Any person who, by the study end date, had received at least 1 doses of an mRNA COVID-19 vaccine at least 90 days following COVID-19 infection or completion of a 2-dose COVID-19 mRNA vaccine series.
<sup>b</sup>The gender variable was not available in the Occupational Health dataset. This was obtained by queries to clinical databases without extracting identifiers. Those without entries in clinical databases were classified as having an unknown gender.

### Cumulative incidence of COVID-19 among boosted and non-boosted individuals with natural and vaccine-induced immunity

Figure 1 compares the cumulative incidence of COVID-19 stratified by immunity type and vaccine boosting status. Among persons with vaccine-mediated immunity, the cumulative incidence of COVID-19 was significantly lower for those boosted compared to those not boosted. However, among those with natural immunity, the cumulative incidence of COVID-19 did not differ between the boosted and the non-boosted in an unadjusted comparison.

**Figure 1.**
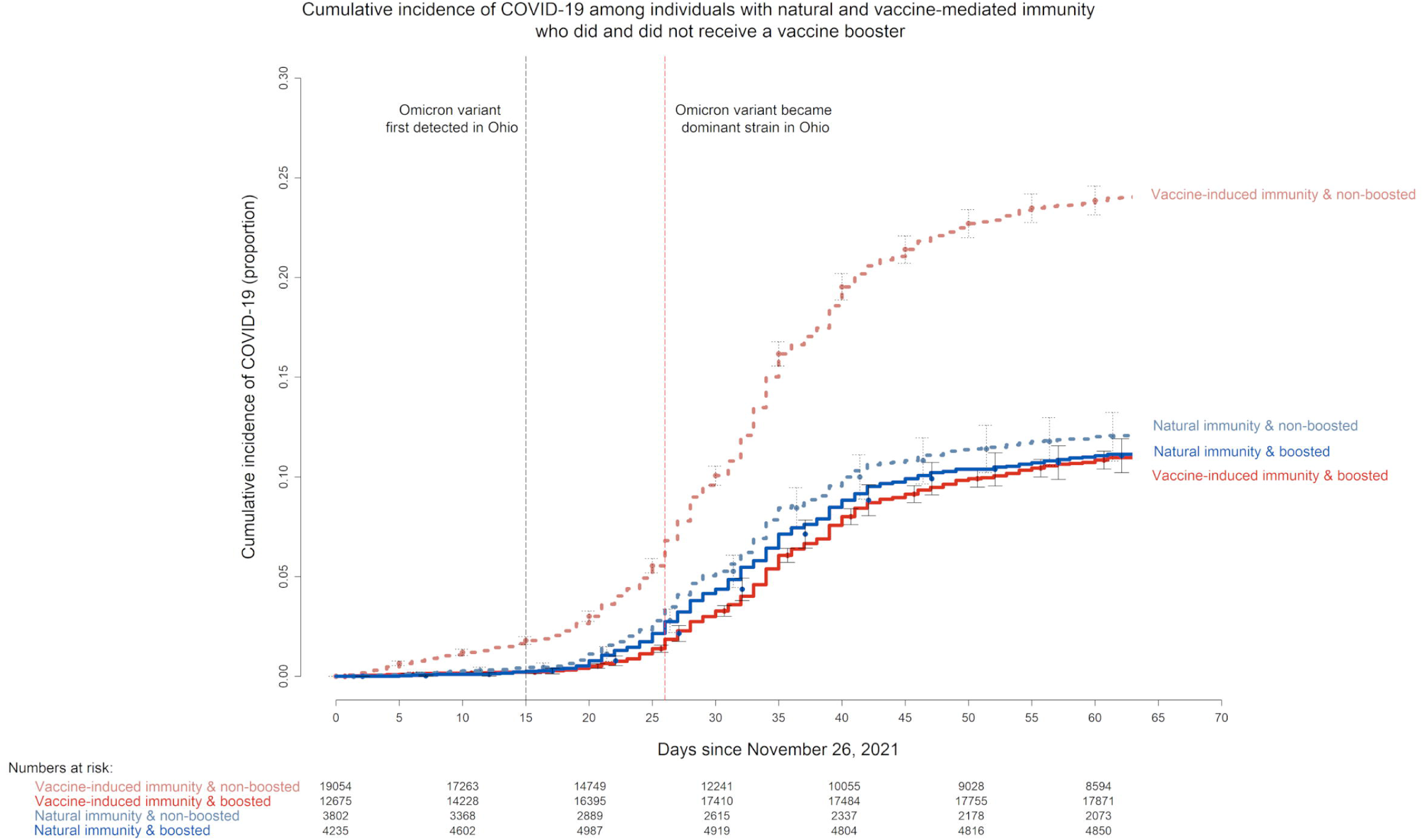
Simon-Makuch plot showing the cumulative incidence of COVID-19 stratified by type of pre-existing immunity (natural or vaccine-induced) and boosting status. Day zero was 26 November 2021, the day the Omicron variant was first detected in the United States. Point estimates and 95% confidence intervals are jittered along the x-axis to improve visibility. Those protected by natural immunity are represented in blue and those protected by vaccine-induced immunity in red. Boosting was a time-dependent covariate whose value changed from “non-boosted” to “boosted” 14 days after receipt of a vaccine booster. Those boosted are represented by bold lines and those who remained non-boosted by dashed lines.

### Time since proximate overt immunologic challenge

Figure 2 shows the risk of COVID-19 stratified by time since POIC, separately for those with natural immunity and those with vaccine-induced immunity.

**Figure 2.**
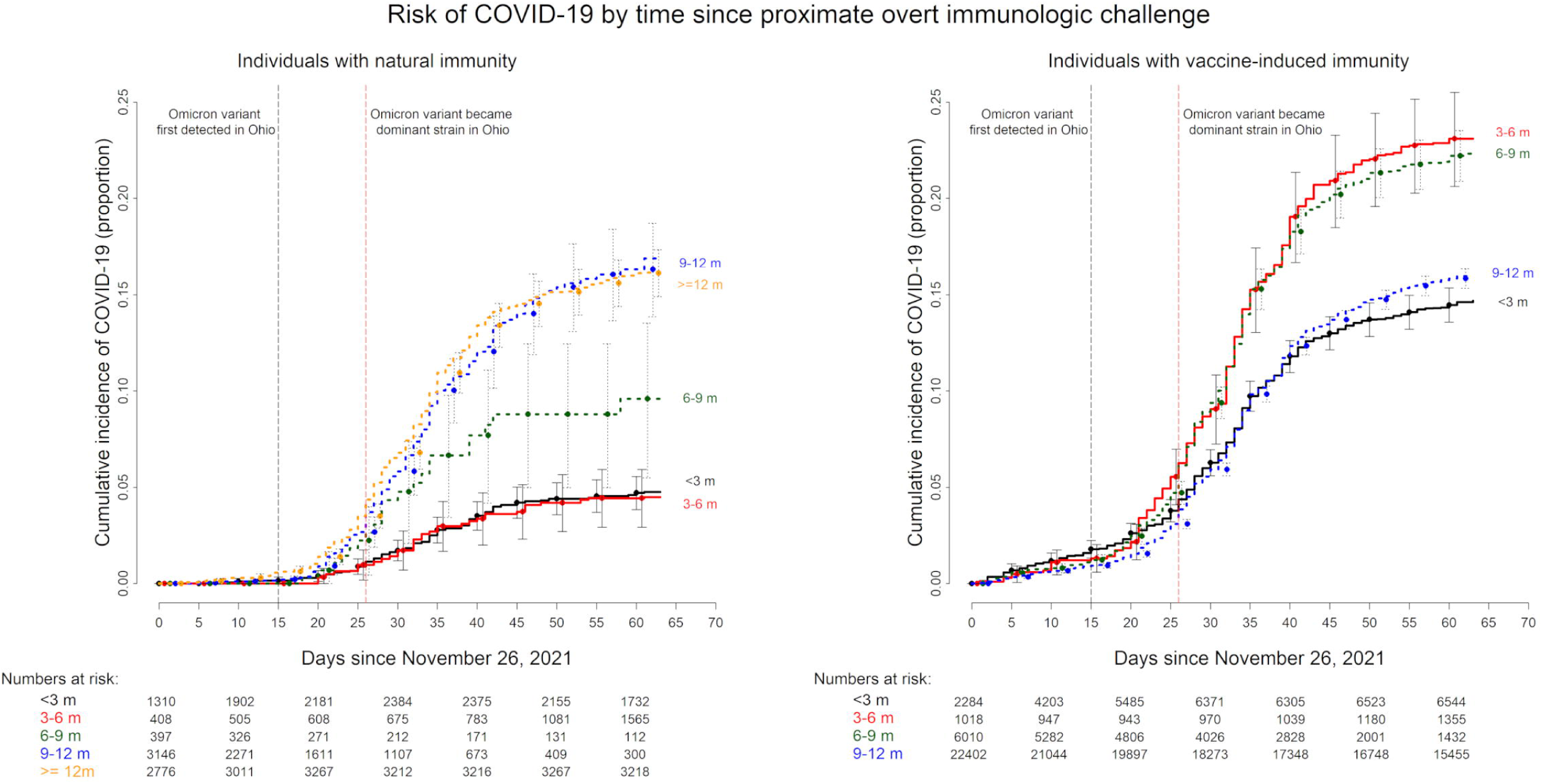
Simon-Makuch plot showing the cumulative incidence of COVID-19 among subjects stratified by time since proximate immune-boosting event (POIC) as a time-dependent covariate. The left panel shows the cumulative incidence for those with natural immunity and the right one for those with vaccine-induced immunity. Day zero was 26 November 2021, the day the Omicron variant was first detected in the United States. Point estimates and 95% confidence intervals are jittered along the x-axis to improve visibility. Receipt of a booster vaccine (as a time-dependent covariate) was considered an immunologic challenge and would result in data for that subject to move to the ‘<3 m’ group on the date of the booster.

For those with natural immunity, the risk of COVID-19 was lowest for POIC within the preceding 6 months. POIC between 6-9 months had a higher risk, and POIC 9 months or longer in the past had an even higher risk. These findings suggest that those with prior COVID-19, caused by a pre-Omicron variant, enjoy a certain level of protection against the Omicron variant for up to 6 months, and that such protection wanes off gradually after 6 months.

For those with vaccine-induced immunity, the risk of COVID-19 was higher for POIC 3-6 or 6-9 months previously compared to POIC within the preceding 3 months, suggesting that the protection against the Omicron variant from two doses of an mRNA vaccine wanes after 3 months. Surprisingly, POIC 9-12 months previously had a lower risk of COVID-19 than POIC 3-9 months previously, and a similar risk to POIC within the preceding 3 months. The explanation for this seeming anomaly is unlikely to be that people transiently lose vaccine-induced immunity after 3 months and regain it after 9 months. This finding could possibly be explained by the fact that those whose POIC occurred 9-12 months previously were those who would have faced the Delta variant within the preceding 3 months with waning vaccine-induced immunity (being past 6 months from their original vaccination) [11], and many of whom may have been inadvertently boosted by an unrecognized asymptomatic or pauci-symptomatic infection with the Delta variant. Those who were vaccinated 3-6 and 6-9 months prior to the start of this study would have been within 6 months of their vaccination during the Delta variant surge, thereby protected from a Delta variant infection at the time [11,16], and thus would not have had the benefit of a boost to their immunity from a Delta variant infection.

### Timing of vaccination after COVID-19

Among persons with natural immunity, in the absence of subsequent vaccination, the risk of COVID-19 was substantially higher for those with prior infection more than 6 months in the past compared to those with prior infection within 6 months (Figure 3). Among those with prior infection 6 months or longer in the past, those vaccinated after COVID-19 had lower risk of COVID-19 than those not. Among those with prior infection within 6 months, protection from vaccination did not statistically significantly differ from the unvaccinated group. A single infection within the <6 months and vaccinated group would make the cumulative incidence of COVID-19 in that group the same as that of the <6 months and unvaccinated group (note the small at risk sample size). Notably, those previously infected within the preceding 6 months and subsequently unvaccinated still had a risk of COVID-19 that was significantly lower than that of those previously infected more than 6 months earlier and subsequently vaccinated. This suggests that natural immunity acquired from an earlier variant of COVID-19 provides substantial protection against the Omicron variant for at least 6 months even in the absence of a vaccine. Six months or later after COVID-19, vaccination provides significant protection.

**Figure 3.**
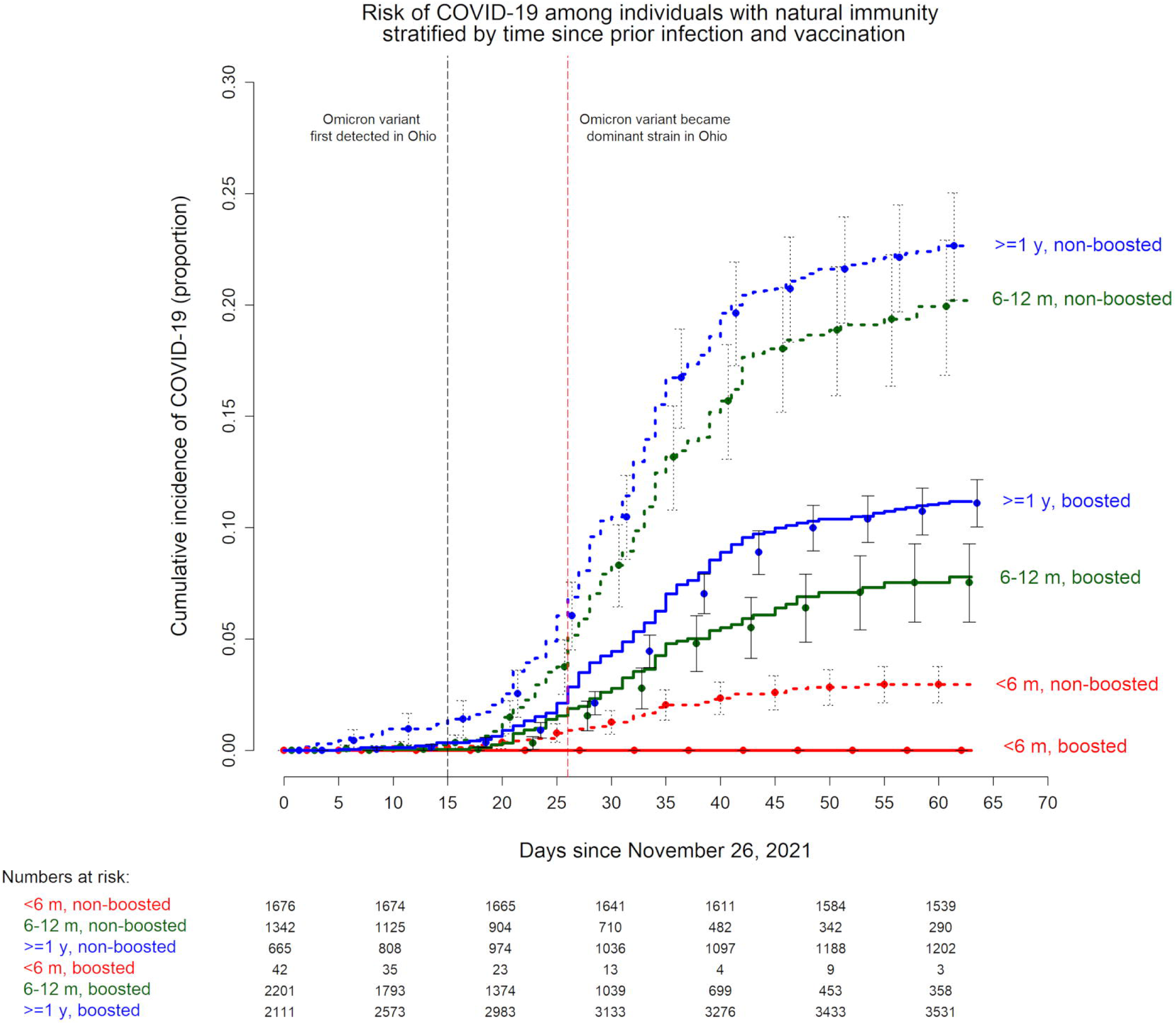
Simon-Makuch plot comparing the cumulative incidence of COVID-19 among subjects with natural immunity from prior infection, stratified by boosting status and time since prior infection. Day zero was 26 November 2021, the day boosters became officially available to healthcare personnel at our institution. Point estimates and 95% confidence intervals are jittered along the x-axis to improve visibility. Time since prior infection strata (as a time-dependent covariate) are represented by different colors. Those boosted (as a time-dependent covariate) are represented by bold lines and those who remained non-boosted by dashed lines.

### Number of vaccine doses after COVID-19

Among individuals with natural immunity from prior infection, those who received 1 dose of vaccine had a significantly lower risk of COVID-19 than those who received no vaccine, but those who received two doses had a higher risk of COVID-19 than those who received a single dose and a risk that was no lower than those who received no vaccine (Figure 4). Those who received 3 doses appeared to have a lower risk than those who received no vaccine, but a higher risk than those who received a single dose.

**Figure 4.**
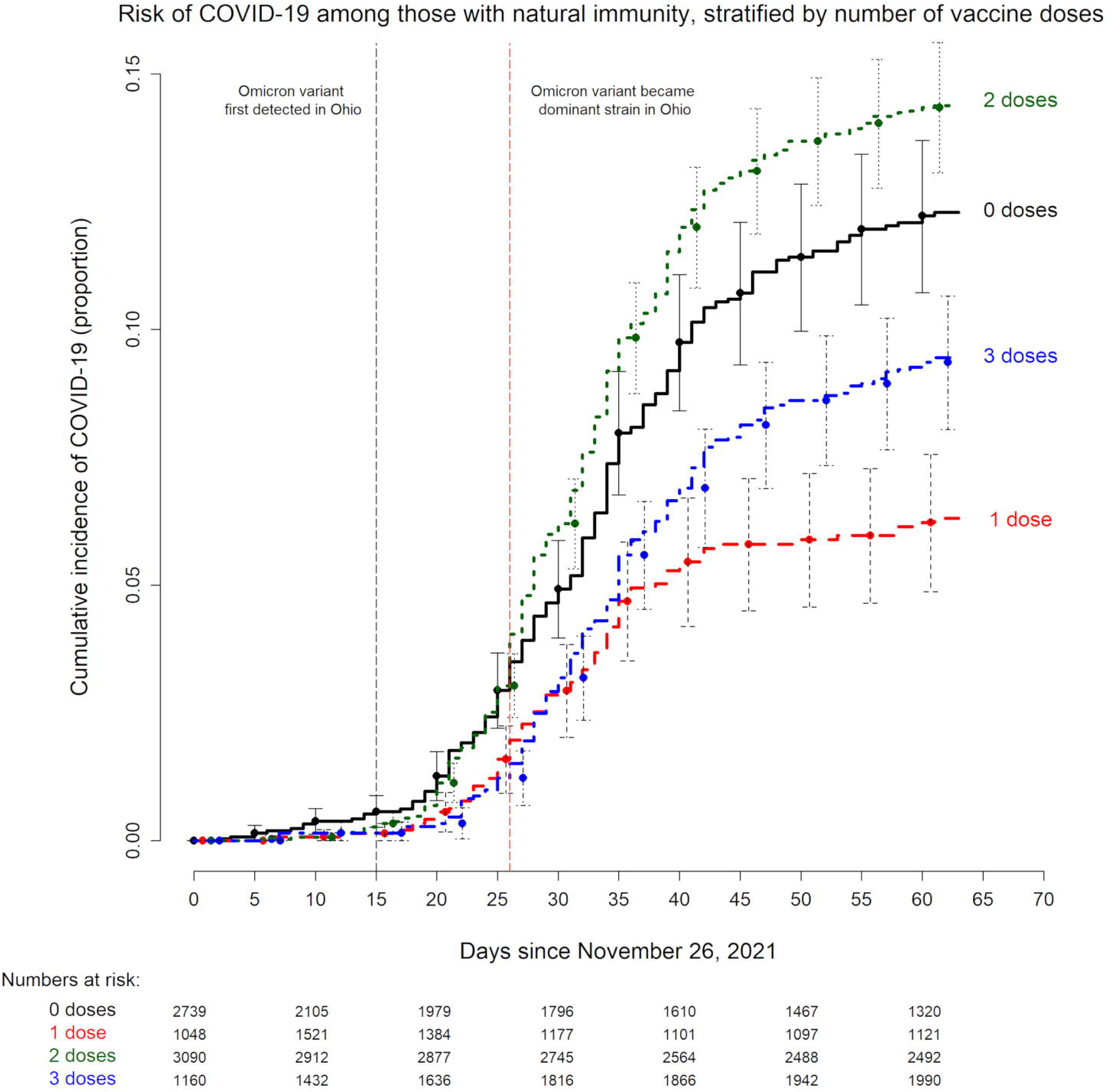
Simon-Makuch plot comparing the cumulative incidence of COVID-19 among individuals with natural immunity from prior infection stratified by number of vaccine doses received (as a time-dependent covariate). Day zero was 26 November 2021, the day boosters became officially available to healthcare personnel at our institution. Point estimates and 95% confidence intervals are jittered along the x-axis to improve visibility.

It is possible that there are unrecognized factors in play here, but this association held in multivariable analysis described later. In the absence of another reasonable explanation, this finding raises the intriguing possibility that a second dose of vaccine given shortly after the first in persons with pre-existing natural immunity might nullify the protection that a single dose of vaccine would otherwise provide.

### Effect of a vaccine booster on occurrence of COVID-19 in multivariable analyses

Boosting with a COVID-19 vaccine designed for an earlier variant was associated with significantly reduced risk of infection with the Omicron variant in multivariable Cox proportional hazards regression analyses, among people previously vaccinated (Table 3) or previously infected for whom it was more than 6 months past their prior infection or vaccination (Table 4).

**Table 3.**
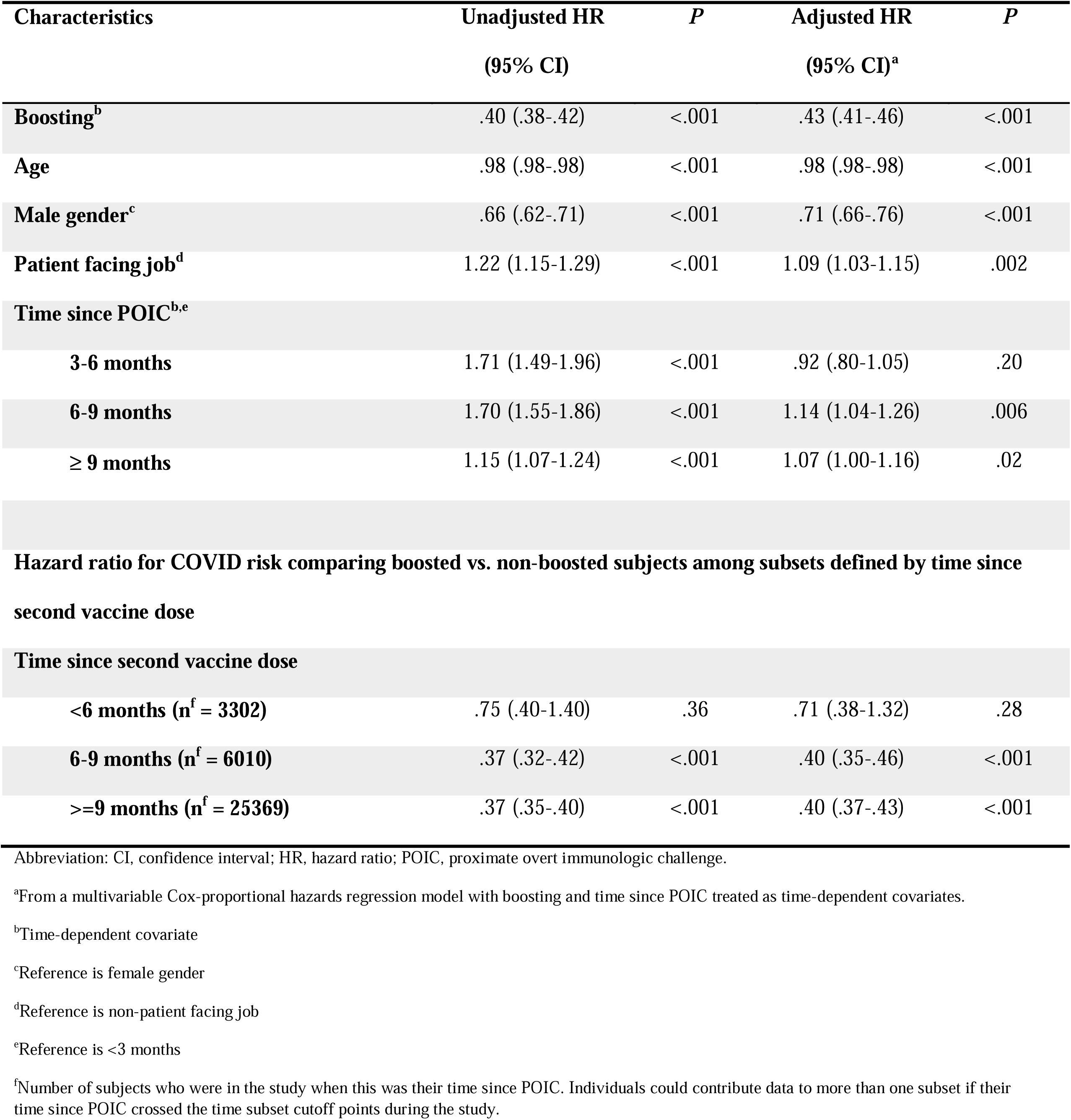
Unadjusted and Adjusted Associations with Time to COVID-19 for Individuals with Vaccine-induced Immunity.

| Characteristics | Unadjusted HR<br>(95% CI) | *P* | Adjusted HR<br>(95% CI)<sup>a</sup> | *P* |
| --- | --- | --- | --- | --- |
| **Boosting<sup>b</sup>** | .40 (.38-.42) | <.001 | .43 (.41-.46) | <.001 |
| **Age** | .98 (.98-.98) | <.001 | .98 (.98-.98) | <.001 |
| **Male gender<sup>c</sup>** | .66 (.62-.71) | <.001 | .71 (.66-.76) | <.001 |
| **Patient facing job<sup>d</sup>** | 1.22 (1.15-1.29) | <.001 | 1.09 (1.03-1.15) | .002 |
| **Time since POIC<sup>b,e</sup>** |  |  |  |  |
| **3-6 months** | 1.71 (1.49-1.96) | <.001 | .92 (.80-1.05) | .20 |
| **6-9 months** | 1.70 (1.55-1.86) | <.001 | 1.14 (1.04-1.26) | .006 |
| **≥ 9 months** | 1.15 (1.07-1.24) | <.001 | 1.07 (1.00-1.16) | .02 |
| **Time since second vaccine dose** |  |  |  |  |
| **<6 months (n<sup>f</sup> = 3302)** | .75 (.40-1.40) | .36 | .71 (.38-1.32) | .28 |
| **6-9 months (n<sup>f</sup> = 6010)** | .37 (.32-.42) | <.001 | .40 (.35-.46) | <.001 |
| **>=9 months (n<sup>f</sup> = 25369)** | .37 (.35-.40) | <.001 | .40 (.37-.43) | <.001 |

Abbreviation: CI, confidence interval; HR, hazard ratio; POIC, proximate overt immunologic challenge.
<sup>a</sup>From a multivariable Cox-proportional hazards regression model with boosting and time since POIC treated as time-dependent covariates.
<sup>b</sup>Time-dependent covariate
<sup>c</sup>Reference is female gender
<sup>d</sup>Reference is non-patient facing job
<sup>e</sup>Reference is <3 months
<sup>f</sup>Number of subjects who were in the study when this was their time since POIC. Individuals could contribute data to more than one subset if their time since POIC crossed the time subset cutoff points during the study.

**Table 4.**
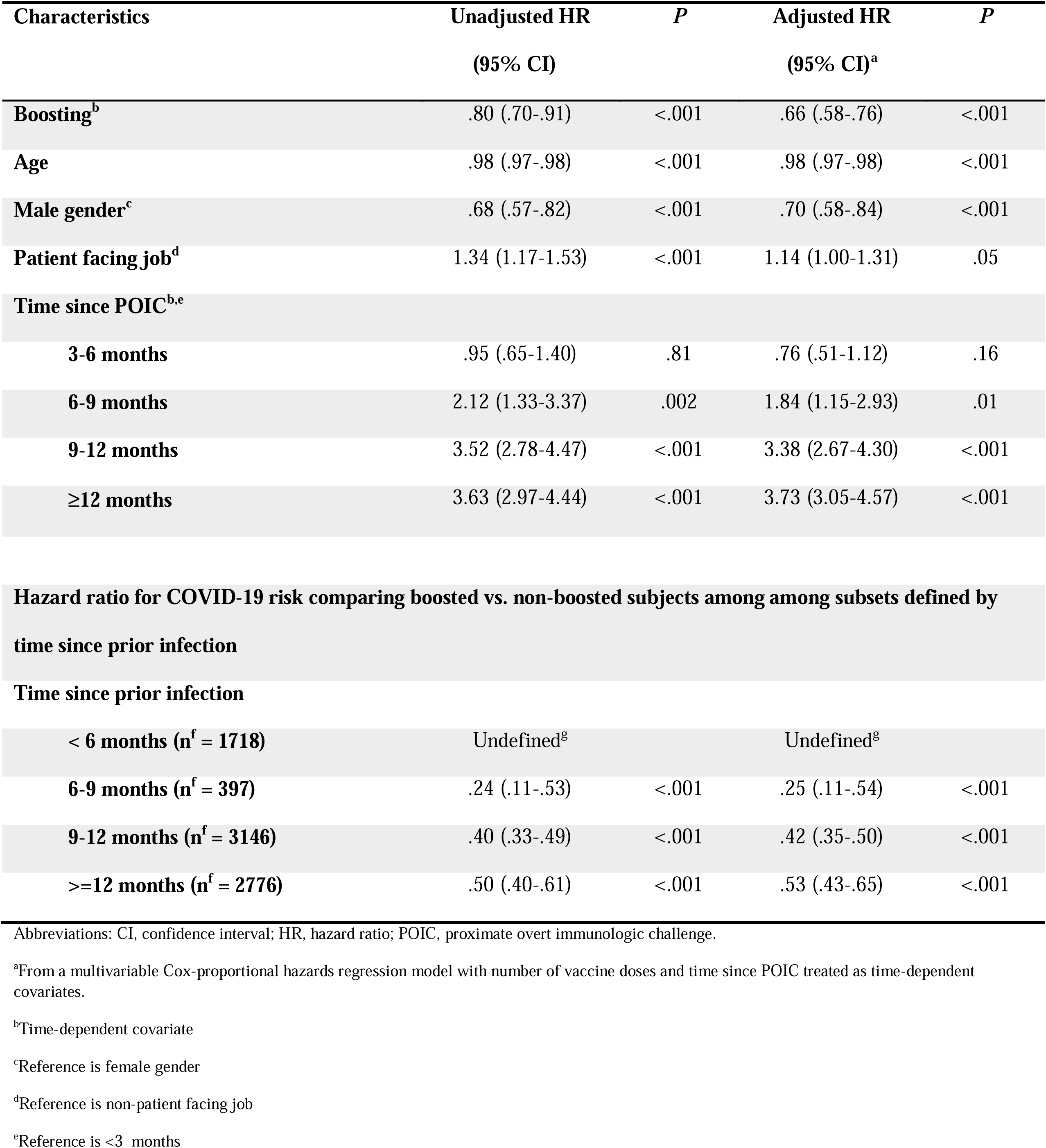
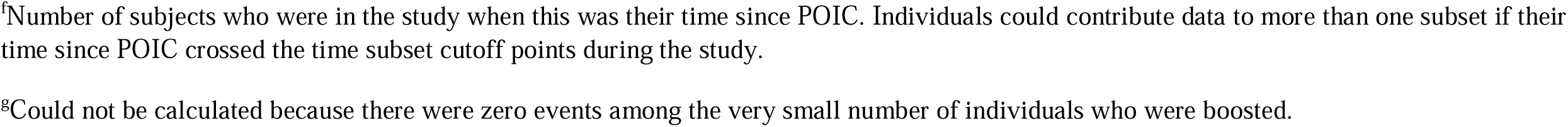
Unadjusted and Adjusted Associations with Time to COVID-19 for Individuals with Natural Immunity from Prior Infection.

| Characteristics | Unadjusted HR<br>(95% CI) | *P* | Adjusted HR<br>(95% CI)<sup>a</sup> | *P* |
| --- | --- | --- | --- | --- |
| **Boosting<sup>b</sup>** | .80 (.70-.91) | <.001 | .66 (.58-.76) | <.001 |
| **Age** | .98 (.97-.98) | <.001 | .98 (.97-.98) | <.001 |
| **Male gender<sup>c</sup>** | .68 (.57-.82) | <.001 | .70 (.58-.84) | <.001 |
| **Patient facing job<sup>d</sup>** | 1.34 (1.17-1.53) | <.001 | 1.14 (1.00-1.31) | .05 |
| **Time since POIC<sup>b,e</sup>** |  |  |  |  |
| **3-6 months** | .95 (.65-1.40) | .81 | .76 (.51-1.12) | .16 |
| **6-9 months** | 2.12 (1.33-3.37) | .002 | 1.84 (1.15-2.93) | .01 |
| **9-12 months** | 3.52 (2.78-4.47) | <.001 | 3.38 (2.67-4.30) | <.001 |
| **≥12 months** | 3.63 (2.97-4.44) | <.001 | 3.73 (3.05-4.57) | <.001 |
| **Time since prior infection** |  |  |  |  |
| **< 6 months (n<sup>f</sup> = 1718)** | Undefined<sup>g</sup> |  | Undefined<sup>g</sup> |  |
| **6-9 months (n<sup>f</sup> = 397)** | .24 (.11-.53) | <.001 | .25 (.11-.54) | <.001 |
| **9-12 months (n<sup>f</sup> = 3146)** | .40 (.33-.49) | <.001 | .42 (.35-.50) | <.001 |
| **>=12 months (n<sup>f</sup> = 2776)** | .50 (.40-.61) | <.001 | .53 (.43-.65) | <.001 |

Abbreviations: CI, confidence interval; HR, hazard ratio; POIC, proximate overt immunologic challenge.
<sup>a</sup>From a multivariable Cox-proportional hazards regression model with number of vaccine doses and time since POIC treated as time-dependent covariates.
<sup>b</sup>Time-dependent covariate
<sup>c</sup>Reference is female gender
<sup>d</sup>Reference is non-patient facing job
<sup>e</sup>Reference is <3 months
<sup>f</sup>Number of subjects who were in the study when this was their time since POIC. Individuals could contribute data to more than one subset if their time since POIC crossed the time subset cutoff points during the study.
<sup>g</sup>Could not be calculated because there were zero events among the very small number of individuals who were boosted.

When the effect of number of vaccine doses was analyzed in multivariable analysis, for individuals with natural immunity, there was no advantage to more than one dose of vaccine, and those who received two doses were at significantly higher risk of getting COVID-19 than those who received a single dose (Table 5), supporting the findings of the unadjusted comparison visually depicted in figure 4.

**Table 5.** Effect of Number of Vaccine Doses on Risk of COVID-19 for Individuals with Natural Immunity from Prior Infection.

| Characteristics | Unadjusted HR<br>(95% CI) | *P* | Adjusted HR<br>(95% CI)<sup>a</sup> | *P* |
| --- | --- | --- | --- | --- |
| **Number of vaccine doses<sup>b,c</sup>** |  |  |  |  |
| **0** | 1.99 (1.54-2.57) | <.001 | 2.44 (1.88-3.15) | <.001 |
| **2** | 2.36 (1.85-3.00) | <.001 | 1.54 (1.21-1.97) | <.001 |
| **3** | 1.52 (1.17-1.98) | .002 | 1.01 (.77-1.32) | .96 |
| **Age** | .98 (.97-.98) | < .001 | .98 (.98-.99) | <.001 |
| **Male gender<sup>d</sup>** | .68 (.57-.81) | < .001 | .73 (.60-.87) | <.001 |
| **Patient facing job<sup>e</sup>** | 1.33 (1.17-1.53) | < .001 | 1.13 (.99-1.30) | .07 |
| **Time since prior infection<sup>b,f</sup>** |  |  |  |  |
| **3-6 months** | 1.97 (1.06-3.66) | .03 | 2.19 (1.28-4.08) | .01 |
| **6-9 months** | 4.14 (2.12-8.08) | <.001 | 4.73 (2.41-9.26) | <.001 |
| **9-12 months** | 7.52 (4.37-12.93) | <.001 | 10.27 (5.92-17.81) | <.001 |
| **≥12 months** | 7.87 (4.63-13.37) | <.001 | 11.29 (6.58-19.40) | <.001 |

Abbreviations: CI, confidence interval; HR, hazard ratio.
<sup>a</sup>From a multivariable Cox-proportional hazards regression model with number of vaccine doses and time since prior infection treated as time-dependent covariates.
<sup>b</sup>Time-dependent covariate
<sup>c</sup>Reference is 1 dose
<sup>d</sup>Reference is female gender
<sup>e</sup>Reference is non-patient facing job
<sup>f</sup>Reference is <3 months

## DISCUSSION

This study corroborates findings from earlier studies that natural immunity from prior infection is more robust than immunity acquired through vaccination [11,17,18], and additionally finds that individuals previously infected with COVID-19 retain substantial protection against the Omicron variant for at least 6 months in the absence of vaccination. For those with waning natural or vaccine-induced immunity (6 months or more after prior infection or vaccination), boosting with a vaccine designed for an earlier variant of COVID-19 still provides significant protection against infection with the Omicron variant. For those with waning natural immunity, a single dose of vaccine provides better protection than multiple doses.

The strengths of our study include its large sample size and a study start date that resulted in all prior infections being pre-Omicron variant infections and the vast predominance of incident infections being Omicron variant infections. Given that this was a study among employees of a health system, that recognized very early the critical importance of maintaining an effective workforce during the pandemic, we had an accurate accounting of who had COVID-19, when they were diagnosed with COVID-19, who received a COVID-19 vaccine, and when they received it. The time-to-event analysis design allowed for important covariates that change over time to be adjusted in a time-dependent manner.

The study has its limitations. Individuals with unrecognized asymptomatic prior infections would have been misclassified as previously uninfected, resulting in underestimating the protective effect of prior infection. Many asymptomatic incident infections were probably missed. There is little reason to suppose, however, that they would have been missed in the various groups at rates disproportionate enough to change the directionality of the study’s findings. Because our employee health symptom-monitoring processes were overwhelmed by disease volume during the Omicron phase of the pandemic, we were unable to distinguish between symptomatic and asymptomatic infections and had to limit our analyses to all detected infections. We did not have a way to adjust for behavioral differences and household exposures, both of which can strongly influence risk of COVID-19. Our study of healthcare personnel included no children and few elderly subjects, and the majority would not have been immunocompromised. Lastly, knowing that the Omicron variant causes milder infection than the Delta variant, the clinical impact of protective effect of vaccine boosting on severe infections would be smaller than the protective effect on infections overall that this study found.

The findings of this study have important implications. Those who have had COVID-19 are better protected against the Omicron variant than those who received two doses of an mRNA COVID-19 vaccine, with protection lasting at least 6 months. There is little to be gained by vaccinating those who are within 6 months of their prior COVID-19 infection. When such individuals are vaccinated to boost waning natural immunity, an intervention to be considered at least 6 months after their prior infection, they should be vaccinated with a single dose of vaccine.

In conclusion, natural immunity from prior COVID-19 provides substantial protection against the Omicron variant for at least 6 months. Boosting individuals with waning immunity, with a COVID-19 vaccine not designed for the Omicron variant protects against Omicron variant infection in both previously vaccinated and previously infected individuals. There is no advantage to administering more than 1 dose of vaccine to previously infected persons. The elderly, children, and the immunocompromised, were not represented or inadequately represented in this study, and caution should be exercised in extrapolating these findings to those populations.

## Data Availability

All data produced in the present study are available upon reasonable request to the authors

## Notes

### Author contributions

N. K. S.: Conceptualization, methodology, validation, investigation, data curation, software, formal analysis, visualization, writing-original draft preparation, writing-reviewing and editing, supervision, project administration. P. S.: Data curation, validation, formal analysis, visualization, writing-reviewing and editing. P. C. B.: Resources, investigation, validation, writing-reviewing and editing. A. S. N.: Methodology, formal analysis, visualization, validation, writing-reviewing and editing.

P. T.: Resources, writing-reviewing and editing. S. M. G.: Project administration, resources, writing-reviewing and editing.

### Potential conflicts of interest

The authors: No reported conflicts of interest. All authors have submitted the ICMJE Form for Disclosure of Potential Conflicts of Interest. Conflicts that the editors consider relevant to the content of the manuscript have been disclosed.

### Funding

None.

## Notes

### Competing Interest Statement

The authors have declared no competing interest.

### Funding Statement

This study did not receive any funding

### Author Declarations

The study was approved by the Cleveland Clinic Institutional Review Board as exempt research (IRB no. 21-1163). A waiver of informed consent and waiver of HIPAA authorization were approved to allow access to de-identified health information by the research team.

